# Rare extreme polygenic risk scores strongly indicate Alzheimer’s disease risk

**DOI:** 10.64898/2026.09.08.26362504

**Authors:** Elizabeth L. Ward, Peter T. Nelson, Yuriko Katsumata, David W. Fardo, Gregory A. Jicha, the Alzheimer’s Disease Neuroimaging Initiative, Justin B. Miller

## Abstract

**Background:** Alzheimer’s disease (AD) pathology often accumulates years before memory loss, creating a need to identify high-risk individuals presymptomatically. Polygenic risk scores (PRS) stratify AD risk, but individual-level predictions remain uncertain. Extreme PRS may identify high-risk individuals independent of *APOE*.

**Methods:** Using GenoPred, extreme tails of 1,752 PRS methods were evaluated across four genetic ancestries using 11,200 autopsy- or clinically-defined AD cases and 19,321 controls (age ≥65) from the AD Sequencing Project (ADSP) Release 5.

**Results:** Individuals in the extreme upper PRS tail were significantly enriched for AD in all ancestries: Admixed American (*P_adj_*=0.02727), African (*P_adj_*=0.004827), East Asian (*P_adj_*=0.02824), and European (*P_adj_*=2.3068×10^−13^). No controls were observed among the 42 European- and 12 African-ancestry individuals with extreme PRS. Extreme PRS spanned *APOE* diplotypes; 38% occurred in non-*APOE* ε4 carriers.

**Conclusions:** Rare subsets of individuals at the most extreme PRS thresholds have markedly elevated AD enrichment across ancestries and *APOE* diplotypes.

**Data Availability:** All data are available through the ADSP, which is managed by The National Institute on Aging Genetics of Alzheimer’s Disease Data Storage Site (NIAGADS).

## Introduction

Alzheimer’s disease (AD) is the most common cause of dementia, affecting over 7.4 million people in the United States and constituting a large and growing public health and economic burden [1]. AD neuropathological changes (ADNC) are understood to begin 15-20 years before the onset of clinical symptoms [2, 3]. This creates a critical window during which presymptomatic individuals could theoretically benefit from early intervention, but only if they can be reliably identified. As such, early risk identification has become a central goal of AD research, particularly for enriching clinical trials with individuals most likely to progress to disease before irreversible neurodegeneration occurs.

AD has a substantial genetic component, with heritability estimates ranging from 60-80% [4]. The *APOE* ε4 allele is the strongest single genetic risk factor, with one copy typically increasing risk ∼3- to 4-fold and two copies increasing risk ∼10- to 15-fold (compared to non-carriers), though effect sizes vary substantially by population ancestry [5, 6]. Beyond *APOE,* large-scale genome-wide association studies (GWAS) have identified more than 75 additional loci contributing to AD risk [7–9], underscoring the polygenic nature of the disease.

Polygenic risk scores (PRS) aggregate the effects of many common variants to estimate an individual’s overall genetic predisposition to AD [10, 11]. They have proven valuable in research settings for predicting age at onset and refining risk stratification among *APOE* ε4 carriers [12], enriching biomarker and trial cohorts [13, 14], and advancing our understanding of polygenic contributions to disease [15]. However, PRS are currently much better calibrated to population-level risk than to individual-level clinical prediction. Although PRS-based enrichment has been shown to improve trial screening efficiency [13, 16], evidence that PRS meaningfully improve individual-level diagnosis or treatment decisions in practice remains limited. Recent reviews have questioned the extent of PRS clinical utility in AD and other complex traits, noting that benefits observed in controlled research settings may not extend robustly to the individual level in practice [17–20]. For AD specifically, *APOE* status often accounts for a large share of genetic risk prediction, and PRS may add only modest incremental value in some populations [21], while accuracy challenges in non-European ancestries further complicate generalizability [22, 23]. Together, these limitations mean that, despite substantial genetic insight, the field still lacks a reliable individual-level tool for identifying high-risk people early enough for presymptomatic intervention or trial enrollment.

Here, we demonstrate pronounced threshold-like behavior in PRS within the AD Sequencing Project (ADSP) Release 5 (R5) cohort, with extreme values strongly enriching for AD case status independent of the *APOE* diplotype, including among *APOE* ε3/ε3 and ε2/ε3 carriers. These results suggest that well-defined extreme PRS thresholds may help identify high-risk individuals before symptom onset and support more efficient clinical trial stratification for AD prevention studies.

## Materials and Methods

All analyses used the ADSP R5 dataset, which provides harmonized whole-genome sequencing (WGS) data spanning 57 multi-ancestry cohorts with substantial representation of African (AFR), Admixed American (AMR), East Asian (EAS), and European (EUR) ancestries [24, 25]. We analyzed 1,752 previously-reported PRS [26] generated using three large-scale commonly-used GWAS from European ancestry (Kunkle, Grenier-Boley [9]; Jansen, Savage [7]; Bellenguez, Küçükali [8]) for each ADSP R5 ancestry using nine algorithms with systematic hyperparameter tuning via the GenoPred [27] pipeline. After reducing redundancy through pairwise correlation analysis, we identified ancestry-specific extreme thresholds that maximized precision for AD case enrichment (requiring ≥10 individuals above threshold) and applied rules-based classification to derive interpretable high-risk strata, both with and without *APOE* (see **Figure 1**).

**Figure 1.**
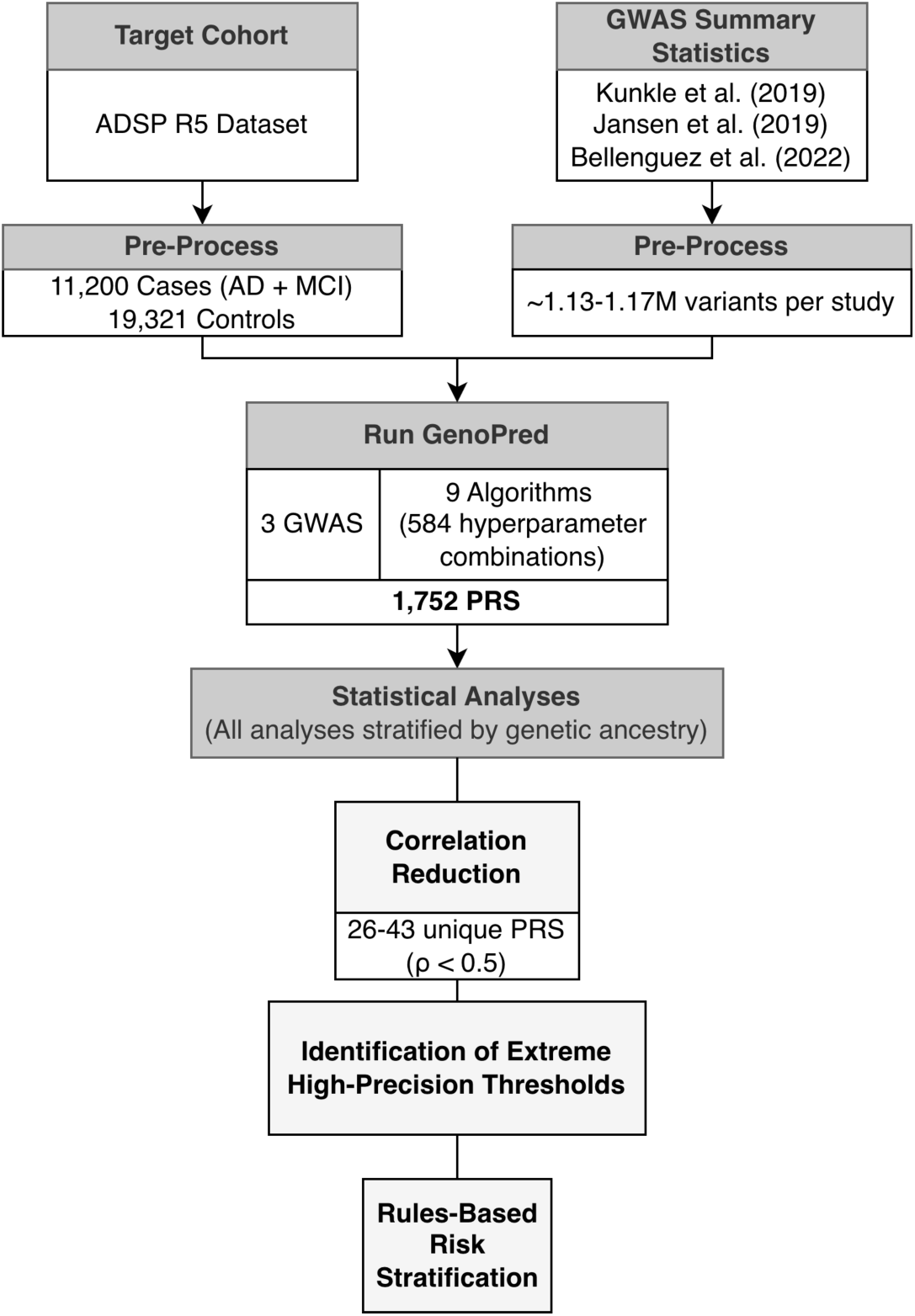
Analytical pipeline for PRS computation and evaluation of high-risk AD thresholds in the ADSP cohort. The process starts with preprocessing the target cohort (left) and three large-scale GWAS summary statistics (right), followed by PRS generation through the GenoPred pipeline, which applies nine algorithms to each of the three GWAS with hyperparameter optimization—producing a total of 1,752 PRS per genetic ancestry. Downstream statistical analyses identify optimal high-risk AD thresholds by maximizing precision and through rules-based risk stratification. All analyses were stratified by genetically inferred ancestry.

### GWAS Summary Statistics

Briefly, PRS were computed using GWAS summary statistics from three major meta-analyses of AD risk, all conducted in predominantly European-ancestry populations.

Kunkle, Grenier-Boley [9] performed an international meta-analysis through the International Genomics of Alzheimer’s Project (IGAP), incorporating consortia including the AD Genetic Consortium (ADGC), European AD Initiative (EADI), Cohorts for Heart and Aging Research in Genomic Epidemiology (CHARGE), and the Genetic and Environmental Risk in AD/Defining Genetic, Polygenic and Environmental Risk for AD Consortium (GERAD/PERADES). Altogether, the meta-analysis comprised 35,274 clinically diagnosed or autopsy-confirmed AD cases and 59,163 controls (total n=94,437).

Jansen, Savage [7] conducted a GWAS-by-proxy (GWAX) by incorporating clinically diagnosed AD with AD-by-proxy phenotypes (based on parental dementia history; genetic correlation r_g_=0.81 with clinical AD), incorporating 71,880 cases and 383,378 controls (total n=455,258) from the Psychiatric Genomics Consortium AD Working Group (PGC-ALZ), ADSP, IGAP, and UK Biobank (UKB). This study reported 29 risk loci enriched in immune-related tissues and amyloid degradation processes.

Bellenguez, Küçükali [8] carried out a two-stage meta-analysis through the European Alzheimer and Dementia Biobank (EADB) consortium, totaling 111,326 clinically diagnosed and proxy AD cases (based on family history reports) and 677,663 controls (total n = 788,989). This identified 75 risk loci (42 novel) associated with amyloid and tau pathways, lipid metabolism, endocytosis, and microglial function.

**Table 1** summarizes key features of the three GWAS/GWAX summary statistics used for PRS calculations.

**Table 1.**
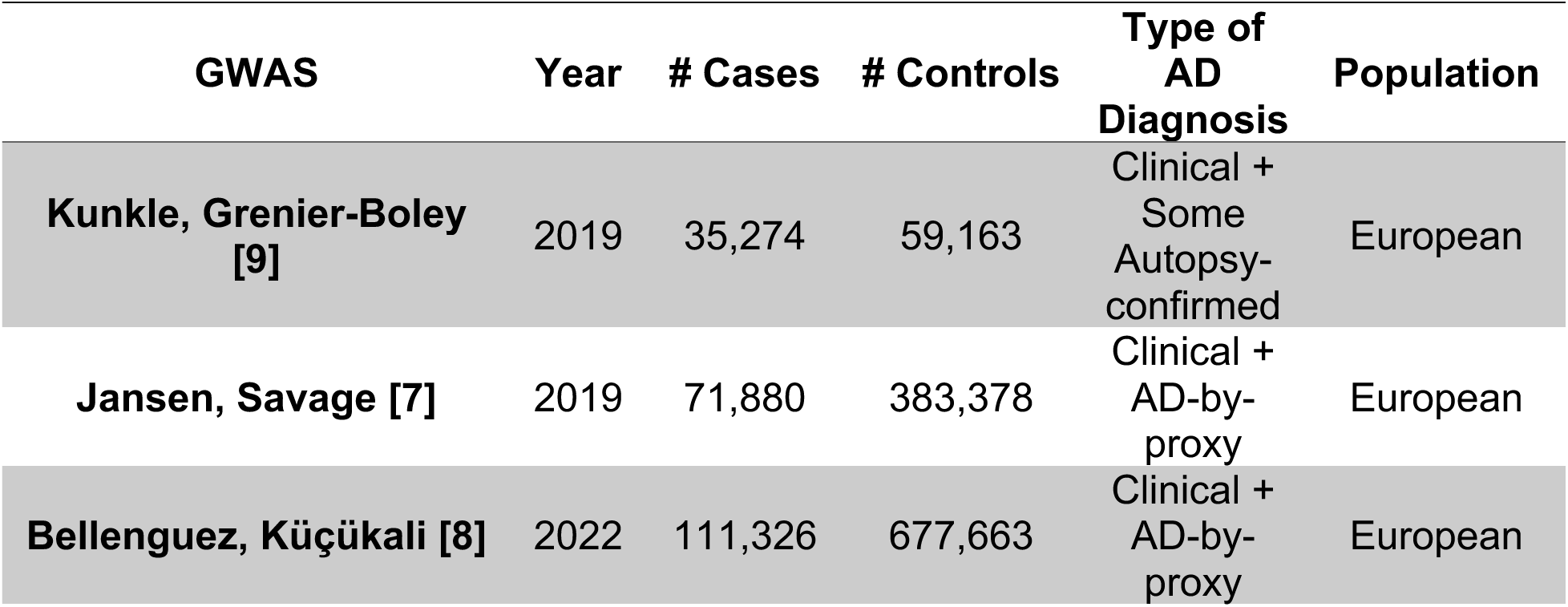
Key characteristics of GWAS summary statistics used in PRS calculations.

| GWAS | Year | # Cases | # Controls | Type of AD Diagnosis | Population |
| --- | --- | --- | --- | --- | --- |
| Kunkle, Grenier-Boley [9] | 2019 | 35,274 | 59,163 | Clinical + Some Autopsy-confirmed | European |
| Jansen, Savage [7] | 2019 | 71,880 | 383,378 | Clinical + AD-by-proxy | European |
| Bellenguez, Küçükali [8] | 2022 | 111,326 | 677,663 | Clinical + AD-by-proxy | European |

### Target cohorts and Sample Selection

We used similar PRS preprocessing and calculation techniques as previously reported [26], and expanded on the analyses to evaluate the association between extreme PRS and AD. Specifically, we used the ADSP R5 dataset for evaluation, which includes data obtained from the AD Neuroimaging Initiative (ADNI) database (adni.loni.usc.edu), since ADNI participants form part of the sequenced and harmonized cohorts in ADSP. The ADNI was launched in 2003 as a public-private partnership, led by Principal Investigator Michael W. Weiner, MD. The primary goal of ADNI has been to test whether serial magnetic resonance imaging (MRI), positron emission tomography (PET), other biological markers, and clinical and neuropsychological assessment can be combined to measure the progression of MCI and early AD.

We extracted sample IDs from the ADSP with confirmed diagnosis of control, MCI, or AD. Exclusions were applied for individuals younger than 65 years at last contact or missing annotations for *APOE* diplotype, age, sex, or race. Additional exclusions removed individuals with non-AD diagnoses: Braak stages 1-6 without an AD diagnosis (n=968), frontotemporal dementia (n=169), Lewy body dementia (n=206), machine learning-assigned cases/controls (n=1,008), Parkinson’s disease (n=11), pathological aging (n=39), progressive supranuclear palsy (n=16), vascular dementia (n=266), or other dementia cases (n=6,426).

Sample IDs from data dictionaries were cross-checked against variant call format (VCF) files, revealing 25 mismatches (**Supplementary Table S1.1**). After review, 21 IDs were corrected to match prior releases, and the remaining four individuals were excluded (**Supplementary Table S1.2**).

Because MCI has a high conversion rate to AD within a few years [28, 29], we combined the MCI and AD groups for all downstream analyses of PRS prediction of AD risk, resulting in 11,200 AD cases and 19,321 controls that passed all filters.

### Preprocessing and Quality Control of the Target Cohort

We applied the standardized GenoPred pipeline [27] for target genotype quality control, relatedness assessment, ancestry inference, harmonization of GWAS summary statistics, PRS computation, and score standardization. GenoPred employs reference panels from the 1000 Genomes Project Phase 3 [30] combined with the Human Genome Diversity Project [31] (1KG+HGDP), restricted to HapMap3 [31] variants, for ancestry inference. By default, it uses European linkage disequilibrium (LD) references as proxy for non-European ancestries due to the limited availability of matched LD panels.

### Relatedness Assessment

Relatedness in the target cohort was evaluated using the KING estimator [32] as implemented in PLINK v1.9 [33], applied to the full sample set. The KING estimator calculates robust pairwise kinship coefficients based on allele-sharing patterns across single nucleotide polymorphisms (SNPs), offering reliable relatedness estimates that remain largely unaffected by population stratification or substructure. To minimize bias, samples with a kinship coefficient exceeding 0.044—corresponding to a fourth-degree relatedness or closer—were excluded from the analysis. All related individuals were removed from subsequent downstream analyses.

### Ancestry Assignment and Within-Population Quality Control

We inferred genetic ancestry using a multinomial elastic-net model trained on the 1KG+HGDP reference panel. Using shared HapMap3 variants that met quality control criteria (minor allele frequency (MAF) >0.05, missingness <0.02, Hardy-Weinberg Equilibrium *P*>1×10^−6^, LD-pruned: window=1000 kb, step=5, r²<0.2), the first six principal components (PCs) were projected onto the target dataset. Assignment to superpopulations was made only when the maximum posterior probability was >95%.

We adhered to GenoPred’s default pipeline for within-population quality control. In populations with at least 100 individuals, 10 within-population PCs were calculated on unrelated samples using LD-pruned variants, and outliers were flagged and removed using k-means clustering based on centroid distances. Because too few individuals remained after filtering, the Central/South Asian (n=39) and Middle Eastern (n=9) ancestry groups were dropped from all downstream PRS analyses. Similarly, individuals who self-identified as Native Hawaiian/Pacific Islander (n=11) or American Indian/Alaska Native (n=78) were excluded due to both limited sample sizes and the lack of well-matched reference populations within the 1KG+HGDP panel. As a result, PRS analyses were restricted to four ancestry groups: AMR, AFR, EAS, and EUR.

### Cohort Composition and Demographics

Following the application of these exclusion criteria, PRS analyses were confined to the four ancestral populations that had sufficient sample sizes: AFR (n = 3,393; 1,260 cases, 2,133 controls), AMR (n = 3,212; 1,304 cases, 1,908 controls), EAS (n = 2,289; 1,018 cases, 1,271 controls), and EUR (n = 13,232; 4,392 cases, 8,840 controls). Other populations and individuals without unclear population projections were removed from the analyses. Demographic characteristics of these cohorts, stratified by ancestry and case-control status, are summarized in **Table 2**.

**Table 2.** Demographics of the final ADSP cohort according to ancestry and case-control status.

| Genetic Ancestry | N | | Female, % | | Age, mean ± SD | | APOE ε4 Carriers, % | |
| --- | --- | --- | --- | --- | --- | --- | --- | --- |
|  | Cases | Controls | Cases | Controls | Cases | Controls | Cases | Controls |
| Admixed American | 1,304 | 1,908 | 64.80 | 67.35 | 75.98 ± 6.55 | 74.09 ± 6.42 | 32.82 | 20.49 |
| African | 1,260 | 2,133 | 72.06 | 77.26 | 76.70 ± 6.77 | 76.20 ± 7.20 | 57.46 | 35.26 |
| East Asian | 1,018 | 1,271 | 60.81 | 48.62 | 75.39 ± 5.65 | 76.57 ± 4.94 | 53.14 | 27.85 |
| European | 4,392 | 8,840 | 54.12 | 59.08 | 76.41 ± 7.65 | 78.61 ± 7.46 | 57.13 | 33.04 |

### Sample- and Variant-Level QC

Initial variant filtering was performed by the ADSP, incorporating FILTER=‘PASS’, VQSR tranche ≥99.7%, read depth ≥10, genotype quality ≥20, call rate >80%, mean depth <500, exclusion of monomorphic variants, and removal of Mendelian inconsistencies. We did not apply any further genotype filters.

PRS calculations were restricted to HapMap3 variants, a common approach that balances computational efficiency with accuracy [34, 35]. GWAS summary statistics were matched to their designated reference genome build, with allele flipping applied where needed, and variants were filtered to exclude those with MAF <0.01, duplicate IDs, invalid p-values, or no variation across samples. After filtering, the following numbers of SNPs were left: 1,160,429 for Bellenguez, Küçükali [8]; 1,129,967 for Jansen, Savage [7], and 1,174,506 for Kunkle, Grenier-Boley [9].

To meet GenoPred’s default requirements (no half-calls or multiallelic sites), ADSP VCFs were first restricted to HapMap3 variants via `bcftools view –regions-file`, then split at multiallelic sites using `bcftools norm -m any`, and finally had half-calls recoded as missing through a combination of `bcftools view` and `awk`. We opted against imputation to avoid introducing potential bias. Additionally, the sample-level QC threshold was relaxed from the default >70% to >60% HapMap3 variants present per individual.

### Polygenic Risk Score Calculation

We assessed AD PRS performance across three GWAS summary statistics and nine algorithms implemented within GenoPred’s standardized framework (DBSLMM [36], Lassosum [37], Lassosum2 [38], LDpred2 [38], MegaPRS [39], PRS-CS [34], PLINK PT+Clump [40], QuickPRS [39], and SBayesRC [41]). Systematic hyperparameter tuning generated 584 PRS per GWAS, for a total of 1,752 PRS across all three.

### Statistical Analyses

#### Identification of Extreme PRS Thresholds

Because we expected a high degree of correlation among similar PRS configurations, we assessed feature correlation of the 1,752 PRS. We set Spearman’s rho (ρ) threshold < 0.5 to establish a reduced set of largely independent signals within each ancestry group. This process resulted in 26-43 independent PRS per ancestry.

For each independent, standardized PRS, we systematically scanned potential thresholds and selected the threshold that maximized precision (i.e., the proportion of AD cases among individuals exceeding the threshold), subject to the constraint that at least 10 individuals in the ancestry group were classified above the threshold.

The PRS configuration (GWAS summary statistics, algorithm, and hyperparameters) yielding the highest precision at its optimal threshold was designated as the ancestry-specific “optimal” extreme PRS. Performance of these extreme thresholds was assessed via chi-square tests comparing the observed distribution of *APOE* diplotypes among individuals exceeding the threshold to the expected AD distribution based on *APOE* diplotypes in the full ancestry cohort. Chi-squared p-values were adjusted by the number of PRS with moderately-low correlation (ρ < 0.5). Results, including the number of independent PRS comparisons informing threshold selection, are summarized in **Table 3**.

**Table 3.**
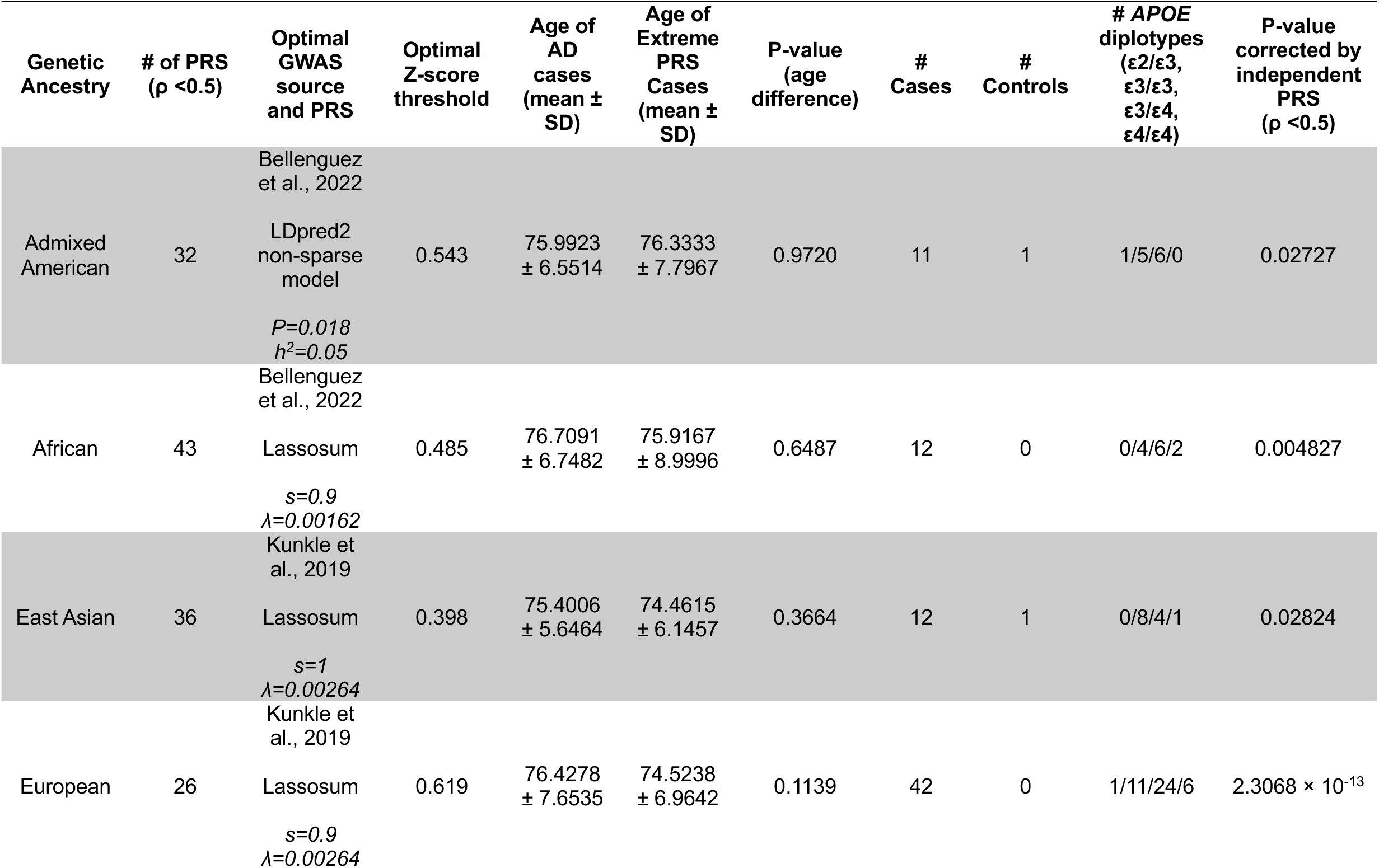
Ancestry-specific optimal extreme PRS thresholds achieving maximum precision for AD case enrichment. For each genetic ancestry group, the table shows the PRS configuration (GWAS source, algorithm, and hyperparameters) and z-score threshold that maximized the proportion of AD cases among individuals exceeding the threshold, subject to the constraint of at least 10 individuals above the threshold. PRS were selected from the set of largely independent scores (ρ<0.5). Age comparisons between all AD cases and those in the extreme PRS stratum are shown, along with the number of cases/controls above threshold, APOE diplotype distribution in the extreme stratum, and chi-square p-value (adjusted for the number of independent PRS tested) comparing observed vs. expected APOE-based AD risk distribution.

To assess potential sample overlap between the ADSP cohort and the GWAS summary statistics used for PRS construction, we compared PRS distributions between likely-overlapped (prior WES participants or ADGC cohort members) and unlikely-overlapped (WGS-only or non-ADGC) subgroups using Mann-Whitney U and Kolmogorov-Smirnov tests with Bonferroni correction. Although statistically significant distributional shifts were detected in EUR and AMR individuals for PRS derived from all three GWAS sources, absolute differences were modest in EUR (maximum KS statistics 0.042-0.015). Larger shifts were observed in AMR (KS 0.25-0.53), while no PRS reached significance in AFR individuals. Excluding likely-overlapped individuals did not change performance rankings or overall conclusions, and the top-performing EUR model retained its rank in both the full and overlap-excluded samples.

### Rules-based Risk Stratification

To determine whether the PRS with unique signals (ρ < 0.5) could be combined into interpretable high-risk thresholds for AD, we applied the JRip algorithm (a Java implementation of RIPPER [42]) in WEKA v3.8.6 [43]. JRip generates compact, ordered sets of human-readable if-then rules using a separate-and-conquer strategy with incremental reduced-error pruning to minimize overfitting and optimize generalization.

A rules-based approach was selected for interpretability and to evaluate each rule as a discrete feature with a fold change and p-value. Since *APOE* has an outsized impact on genetic predisposition for AD, the JRip algorithm was run twice: once including *APOE* and once not including *APOE*. The intent of this analysis is to establish PRS thresholds that result in the highest genetic risk for AD, independent of age or other covariates. Thus, we included only the unique PRS, sex, and (optionally) *APOE* diplotype in the classification algorithm. Default JRip parameters were used.

## Results

### PRS omputation

We computed a total of 1,752 PRS per ancestry group using summary statistics from three GWAS combined with nine algorithms and extensive hyperparameter tuning. PRS were restricted to harmonized HapMap3 variants, retaining ∼1.13-1.17 million SNPs per GWAS source. GenoPred’s reference-based standardization ensured approximately normal distributions with mean z-score ≈ 0 and SD ≈1 within each ancestry, with slight variance reductions expected in non-EUR groups due to European-centric GWAS training.

### Identification of Optimal Extreme PRS Thresholds

For each largely independent PRS within an ancestry group (ρ<0.5), thresholds were systematically evaluated to identify the value that maximized precision (proportion of AD cases among individuals exceeding the threshold) while ensuring at least 10 individuals in the ancestry group were classified above the threshold. The PRS and threshold yielding the highest precision under this constraint was designated as the ancestry-specific optimal extreme PRS.

Ancestry-specific optimal thresholds and performance metrics are detailed in **Table 3**. Notably, precision approached or reached 1.0 in all ancestries, indicating strong enrichment of AD cases in the high-risk strata. Chi-square tests confirmed significant deviation from the expected *APOE*-based AD distribution, with all associations surviving p-value adjustment across the number of largely independent PRS tested per ancestry. The presence of multiple ε3/ε3 cases in high-risk strata further suggests contributions from polygenic signal beyond *APOE*.

Lassosum models were optimal in three ancestries, with perfect precision (no controls above threshold) achieved in AFR and EUR despite the ≥10 individuals coverage requirement. These high-risk thresholds informed the PRS features used in subsequent JRip rules-based stratification.

To contextualize these thresholds within the empirical score distributions, we computed IQR-based statistical outlier bounds for the standardized PRS selected for each ancestry. The upper outlier bounds were: AFR = 0.480 (13 individuals above: 12 cases, 1 control), EUR = 0.5955 (54 individuals above: 49 cases, 5 controls), AMR = 0.5755 (5 individuals above: 5 cases, 0 controls), and EAS = 0.4525 (10 individuals above: 9 cases, 1 control). Notably, optimal precision-maximizing thresholds were at or above the IQR-defined outlier boundary in AFR and EUR groups, indicating that the most discriminating scores in these ancestries represent true statistical extremes. In contrast, optimal thresholds for EAS and AMR groups fell below their respective IQR bounds, suggesting that meaningful case enrichment in these ancestries begins at scores that are elevated but not yet distributional outliers.

### Stratification of AD risk by standardized PRS and *APOE* diplotype across ancestries

To illustrate the interaction between polygenic burden and *APOE* diplotype, we plotted the percentage of individuals with an AD diagnosis for each standardized PRS decile, stratified by *APOE* diplotype, separately for each ancestry group (**Figure 2**). For visualization, we selected—within each ancestry—the single PRS from the reduced set of largely independent PRS (ρ<0.5; **Table 3**) that maximized precision at its extreme high-risk threshold (≥10 individuals above threshold). The AMR control with an extreme PRS was age 90+, while the EAS control with extreme PRS was age 70-75.

**Figure 2.**
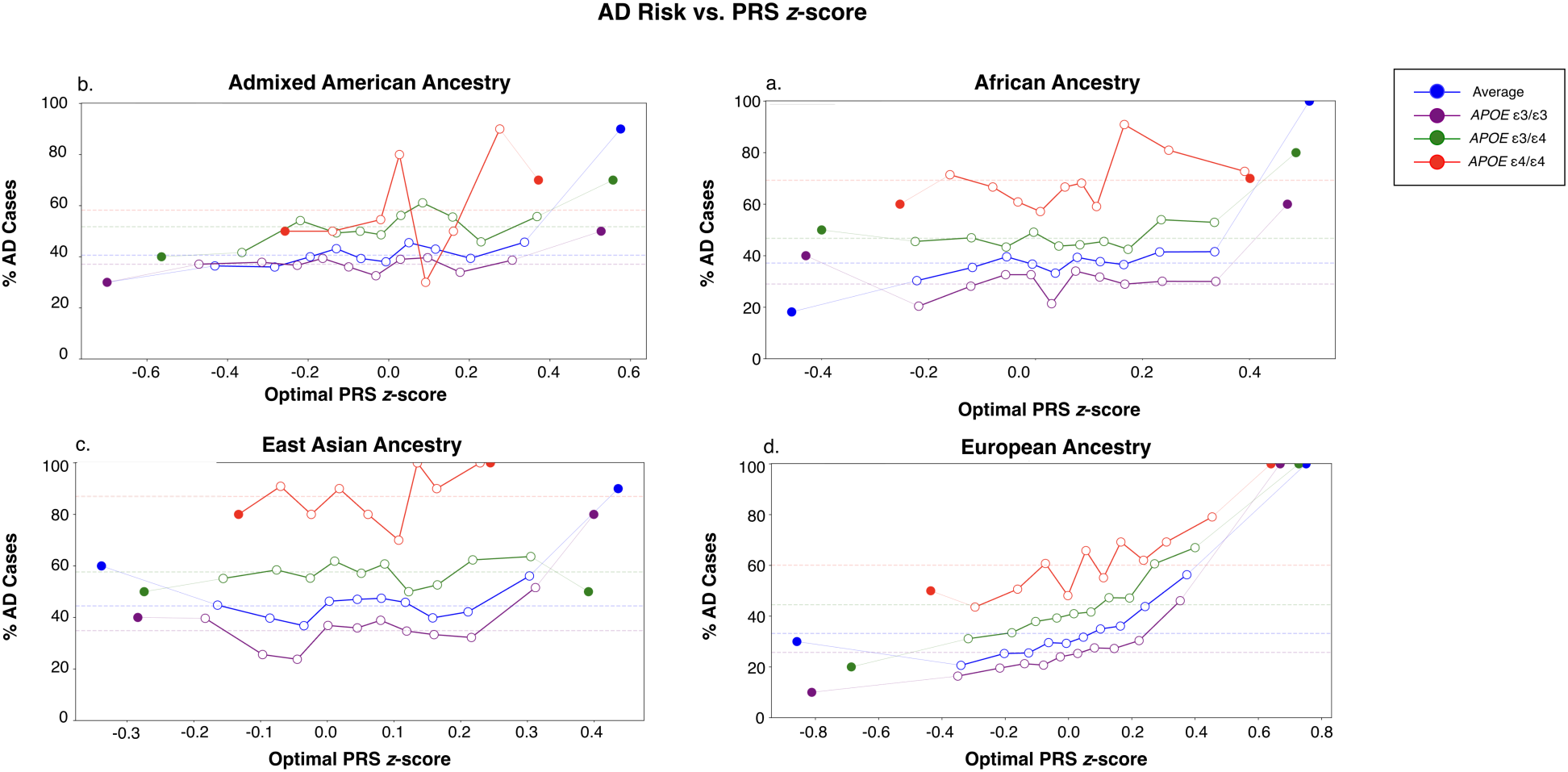
AD prevalence stratified by PRS decile and APOE diplotype across genetic ancestries. Each point represents the **observed AD prevalence within a decile of standardized PRS, plotted separately by APOE diplotype for individuals of (a) AFR, (b) AMR, (c) EAS, and (d) EUR ancestries.** Solid points at either end represent the top and bottom 10 individuals by PRS within each diplotype, rather than a decile. Dashed horizontal lines indicate the overall population-average AD risk conferred by each APOE diplotype alone. For each ancestry, the PRS with the highest precision at its extreme threshold (≥10 individuals) was selected for display. At high PRS extremes, risk approached 80-100% and was largely independent of the APOE diplotype, including among carriers of protective alleles. The selected high-PRS thresholds showed exceptional precision: EUR (threshold ≥0.619; 42 cases, 0 controls), EAS (0.398; 12 cases, 1 control), AMR (0.543; 11 cases, 1 control), and AFR (0.485; 12 cases, 0 controls).

Across non-European ancestries, AD prevalence at low-to-moderate PRS levels broadly aligned with baseline expectations from *APOE* diplotype alone (dashed horizontal lines). At the extreme high-PRS tail, however, prevalence rose sharply, most notably among *APOE* ε3/ε3 carriers, approaching levels typically seen only in *APOE* ε4 carriers.

### Rules-based Analysis of PRS Combinations

The JRip algorithm was applied to the 26-43 largely independent PRS (ρ<0.5), sex, and (in separate iterations) *APOE* diplotype to generate compact, interpretable if-then rules for high-risk AD strata. When *APOE* was included, JRip produced three rules in EUR, six in AFR, one in AMR, and four in EAS (**Supplementary Tables S1-S4**). These rules revealed synergistic effects: several ε3/ε4 + PRS combinations achieved case enrichment comparable to or exceeding ε4/ε4 homozygotes. As shown in **Figure 3** (**Panel A**), the highest-enrichment rules produced AD case proportions of 57.5% (OR 1.29 vs ε3/ε4 baseline, *P*=2.21×10^−15^), up to 80.9% in AFR (OR 1.73 vs ε3/ε4 baseline, *P=*8.82×10^−6^), and 64.9% in EAS (OR 1.13 vs. ε3/ε4 baseline, *P*=0.0026). In AFR, multiple ε3/ε4-based rules surpassed the 69.3% case rate observed in ε4/ε4 carriers. In contrast, AMR showed minimal additional stratification beyond APOE ε3/ε4 status alone.

**Figure 3.**
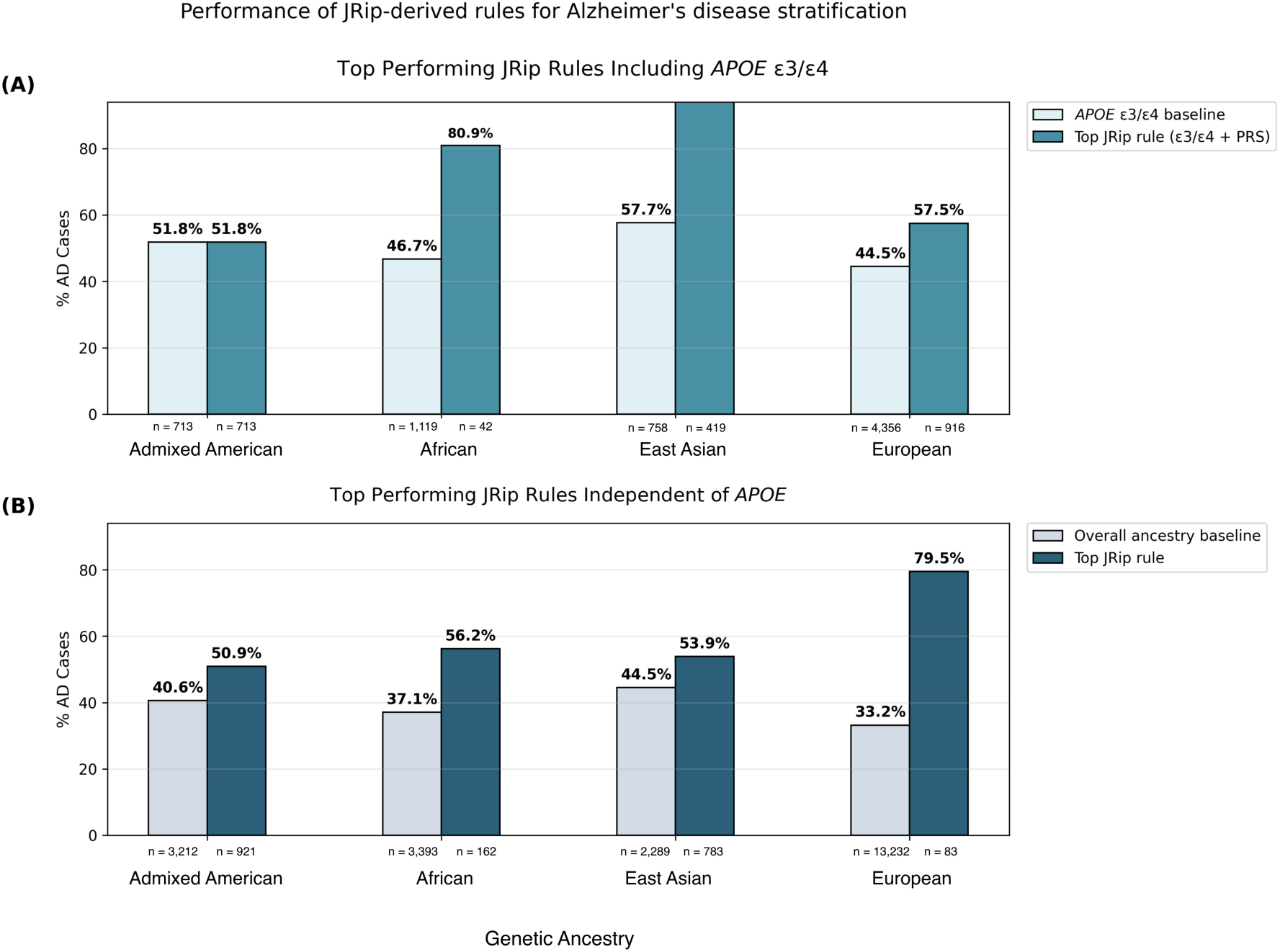
Performance of JRip-derived high-risk classification rules for stratifying AD cases by genetic ancestry. Bars show observed percentage of AD cases for baseline groups versus the top-performing JRip rule in each ancestry group: EUR, AFR, AMR, and EAS. **Panel A:** Rules incorporating APOE e3/e4 genotype (with various PRS algorithms and GWAS sources); for the AMR group, only the baseline APOE e3/e4 rule was produced (no PRS), yielding identical percentages. **Panel B:** APOE-independent rules (using PRS and/or sex only). Data are from Supplementary Tables S1-S8; only the single highest case-enrichment rule per category is shown for clarity. All differences between rules and baselines are statistically significant (details in supplementary tables).+

When *APOE* was excluded (**Supplementary Tables S5-S8**), JRip identified nine rules in EUR, three in AFR, and one each in AMR and EAS. The strongest non-*APOE* signal emerged in EUR, where the top rules achieved 79.5–80.8% AD cases (OR 2.40– 2.43 vs. overall ancestry baseline, *P*≤3.2×10⁻¹³) using only PRS thresholds (**Figure 3, Panel B**). AFR rules reached 56.2% cases (OR 1.51, *P*≤5.3×10⁻⁷), while AMR and EAS showed more modest gains (50.9% and 53.9% cases; OR 1.25 and 1.21, respectively; *P*≤1.76×10⁻⁷). MegaPRS and Lassosum models appeared recurrently across ancestries and rule sets.

## Discussion

This study provides evidence that a small subset of individuals at the extreme tail of ancestry-specific polygenic risk score (PRS) distributions can be identified with near-perfect or perfect precision for AD case status. These high-risk strata were observed independently of the *APOE* diplotype, including among *APOE* ε3/ε3 and ε2/ε3 carriers, which are generally considered normal or low-risk diplotypes. These findings demonstrate that extreme PRS thresholds can identify individuals at exceptionally elevated genetic risk for AD, complementing *APOE* diplotyping as a tool for presymptomatic risk identification.

The extreme-tail precision was not explained by a simple linear extension of PRS-associated risk. Below ancestry-specific cutoffs, PRS provided little additional discrimination beyond *APOE* diplotyping, whereas there was extreme AD risk observed above the extreme cutoffs. This threshold-like behavior suggests that rare combinations of common variants can, in aggregate, define a substantially enriched risk stratum comparable in magnitude to that seen with high-penetrance monogenic variants in genes such as *APP, PSEN1,* or *PSEN2* [44, 45], even though the underlying biological mechanisms differ.

Rules-based analyses further illustrate how these signals could be translated into more generalizable risk strata that extend beyond the extreme tail. Specific combinations of extreme PRS thresholds and *APOE* diplotypes—particularly within the ε3/ε4 diplotype—produced case enrichment comparable to or exceeding the average AD risk for individuals with the ε4/ε4 diplotype in African, East Asian, and European ancestries, while PRS-only rules identified high-risk groups independent of *APOE.* Because these thresholds identify a small, high-confidence subset of individuals that are diagnosed with AD, the thresholds are well suited to enriching biomarker studies and prevention trials with genetically high-risk pre-symptomatic individuals, including candidates not identified through *APOE* diplotyping alone.

We systematically evaluated a large number of PRS configurations (1,752 per ancestry) and identified ancestry-specific thresholds that maximized case enrichment while including at least 10 individuals. This approach underscores that the predictive value of PRS for AD is disproportionately concentrated at the upper tail of the distribution, rather than distributed evenly across it. Notably, extreme PRS remained informative across multiple ancestries despite the use of predominantly European-derived GWAS summary statistics, although performance was attenuated in non-European groups. To contextualize these thresholds within the empirical score distributions, IQR-based outlier bounds were computed for each ancestry; optimal thresholds in AFR and EUR fell at or above these bounds, whereas thresholds in EAS and AMR fell below them. It is worth noting that these more permissive thresholds still achieved strong case enrichment, underscoring that precision-maximizing threshold selection may identify high-risk strata that IQR-based outlier definitions alone would obscure.

Although the AMR control with extreme PRS was age 90+, they had a PRS that was close to the extreme threshold (0.561 observed PRS compared to the 0.543 threshold). The maximum observed PRS value in the AMR cohort was 0.77, indicating that higher scores within the extreme threshold might also have higher confidence.

The EAS control with extreme PRS, on the other hand, was age 70-75 and had the second highest observed PRS in the EAS cohort (0.466 compared to an extreme threshold of 0.398 and a maximum PRS in the EAS cohort of 0.494). Without additional follow-up and longitudinal observation, it is difficult to determine if that individual should be considered a false positive or if they will develop AD later in life.

Several considerations will be important for future work. Due to the relative rarity of individuals with extreme PRS in the ADSP, it is difficult to ascertain the prevalence of extreme AD PRS and their applicability to a more general population. Replication in independent, prospectively determined cohorts will also be needed to establish stable thresholds suitable for trial screening applications. Because PRS can be computed once from genotype data alone, without the recurring cost or participant burden of biomarker or imaging-based screening, extreme PRS thresholds may offer a particularly scalable first-pass enrichment strategy for pre-symptomatic populations, complementing existing approaches based on age, *APOE,* or biomarker status and supporting more efficient evaluation of emerging disease-modifying therapies.

Ultimately, the concentration of high-precision genetic signal at the extreme upper tail of the PRS distribution offers a practical, *APOE-*independent framework for identifying individuals at markedly elevated AD risk, with direct potential application to presymptomatic trial enrichment and target study design in AD research.

## Acknowledgements

The Alzheimer’s Disease Sequencing Project (ADSP) is comprised of two AD genetics consortia and three National Human Genome Research Institute (NHGRI) funded Large Scale Sequencing and Analysis Centers (LSAC). The two AD genetics consortia are the Alzheimer’s Disease Genetics Consortium (ADGC) funded by NIA (U01 AG032984), and the Cohorts for Heart and Aging Research in Genomic Epidemiology (CHARGE) funded by NIA (R01 AG033193), the National Heart, Lung, and Blood Institute (NHLBI), other National Institute of Health (NIH) institutes and other foreign governmental and non-governmental organizations. The Discovery Phase analysis of sequence data is supported through UF1AG047133 (to Drs. Schellenberg, Farrer, Pericak-Vance, Mayeux, and Haines); U01AG049505 to Dr. Seshadri; U01AG049506 to Dr. Boerwinkle; U01AG049507 to Dr. Wijsman; and U01AG049508 to Dr. Goate and the Discovery Extension Phase analysis is supported through U01AG052411 to Dr. Goate, U01AG052410 to Dr. Pericak-Vance and U01 AG052409 to Drs. Seshadri and Fornage.

Sequencing for the Follow Up Study (FUS) is supported through U01AG057659 (to Drs. PericakVance, Mayeux, and Vardarajan) and U01AG062943 (to Drs. Pericak-Vance and Mayeux). Data generation and harmonization in the Follow-up Phase is supported by U54AG052427 (to Drs. Schellenberg and Wang). The FUS Phase analysis of sequence data is supported through U01AG058589 (to Drs. Destefano, Boerwinkle, De Jager, Fornage, Seshadri, and Wijsman), U01AG058654 (to Drs. Haines, Bush, Farrer, Martin, and Pericak-Vance), U01AG058635 (to Dr. Goate), RF1AG058066 (to Drs. Haines, Pericak-Vance, and Scott), RF1AG057519 (to Drs. Farrer and Jun), R01AG048927 (to Dr. Farrer), and RF1AG054074 (to Drs. Pericak-Vance and Beecham).

The ADGC cohorts include: Adult Changes in Thought (ACT) (U01 AG006781, U19 AG066567), the Alzheimer’s Disease Research Centers (ADRC) (P30 AG062429, P30 AG066468, P30 AG062421, P30 AG066509, P30 AG066514, P30 AG066530, P30 AG066507, P30 AG066444, P30 AG066518, P30 AG066512, P30 AG066462, P30 AG072979, P30 AG072972, P30 AG072976, P30 AG072975, P30 AG072978, P30 AG072977, P30 AG066519, P30 AG062677, P30 AG079280, P30 AG062422, P30 AG066511, P30 AG072946, P30 AG062715, P30 AG072973, P30 AG066506, P30 AG066508, P30 AG066515, P30 AG072947, P30 AG072931, P30 AG066546, P20 AG068024, P20 AG068053, P20 AG068077, P20 AG068082, P30 AG072958, P30 AG072959), the Chicago Health and Aging Project (CHAP) (R01 AG11101, RC4 AG039085, K23 AG030944), Indiana Memory and Aging Study (IMAS) (R01 AG019771), Indianapolis Ibadan (R01 AG009956, P30 AG010133), the Memory and Aging Project (MAP) ( R01 AG17917), Mayo Clinic (MAYO) (R01 AG032990, U01 AG046139, R01 NS080820, RF1 AG051504, P50 AG016574), Mayo Parkinson’s Disease controls (NS039764, NS071674, 5RC2HG005605), University of Miami (R01 AG027944, R01 AG028786, R01 AG019085, IIRG09133827, A2011048), the Multi-Institutional Research in Alzheimer’s Genetic Epidemiology Study (MIRAGE) (R01 AG09029, R01 AG025259), the National Centralized Repository for Alzheimer’s Disease and Related Dementias (NCRAD) (U24 AG021886), the National Institute on Aging Late Onset Alzheimer’s Disease Family Study (NIA-LOAD) (U24 AG056270), the Religious Orders Study (ROS) (P30 AG10161, R01 AG15819), the Texas Alzheimer’s Research and Care Consortium (TARCC) (funded by the Darrell K Royal Texas Alzheimer’s Initiative), Vanderbilt University/Case Western Reserve University (VAN/CWRU) (R01 AG019757, R01 AG021547, R01 AG027944, R01 AG028786, P01 NS026630, and Alzheimer’s Association), the Washington Heights-Inwood Columbia Aging Project (WHICAP) (RF1 AG054023), the University of Washington Families (VA Research Merit Grant, NIA: P50AG005136, R01AG041797, NINDS: R01NS069719), the Columbia University Hispanic Estudio Familiar de Influencia Genetica de Alzheimer (EFIGA) (RF1 AG015473), the University of Toronto (UT) (funded by Wellcome Trust, Medical Research Council, Canadian Institutes of Health Research), and Genetic Differences (GD) (R01 AG007584). The CHARGE cohorts are supported in part by National Heart, Lung, and Blood Institute (NHLBI) infrastructure grant HL105756 (Psaty), RC2HL102419 (Boerwinkle) and the neurology working group is supported by the National Institute on Aging (NIA) R01 grant AG033193.

The CHARGE cohorts participating in the ADSP include the following: Austrian Stroke Prevention Study (ASPS), ASPS-Family study, and the Prospective Dementia Registry-Austria (ASPS/PRODEM-Aus), the Atherosclerosis Risk in Communities (ARIC) Study, the Cardiovascular Health Study (CHS), the Erasmus Rucphen Family Study (ERF), the Framingham Heart Study (FHS), and the Rotterdam Study (RS). ASPS is funded by the Austrian Science Fond (FWF) grant number P20545-P05 and P13180 and the Medical University of Graz. The ASPS-Fam is funded by the Austrian Science Fund (FWF) project I904), the EU Joint Programme – Neurodegenerative Disease Research (JPND) in frame of the BRIDGET project (Austria, Ministry of Science) and the Medical University of Graz and the Steiermärkische Krankenanstalten Gesellschaft. PRODEM-Austria is supported by the Austrian Research Promotion agency (FFG) (Project No. 827462) and by the Austrian National Bank (Anniversary Fund, project 15435. ARIC research is carried out as a collaborative study supported by NHLBI contracts (HHSN268201100005C, HHSN268201100006C, HHSN268201100007C, HHSN268201100008C, HHSN268201100009C, HHSN268201100010C, HHSN268201100011C, and HHSN268201100012C). Neurocognitive data in ARIC is collected by U01 2U01HL096812, 2U01HL096814, 2U01HL096899, 2U01HL096902, 2U01HL096917 from the NIH (NHLBI, NINDS, NIA and NIDCD), and with previous brain MRI examinations funded by R01-HL70825 from the NHLBI. CHS research was supported by contracts HHSN268201200036C, HHSN268200800007C, N01HC55222, N01HC85079, N01HC85080, N01HC85081, N01HC85082, N01HC85083, N01HC85086, and grants U01HL080295 and U01HL130114 from the NHLBI with additional contribution from the National Institute of Neurological Disorders and Stroke (NINDS). Additional support was provided by R01AG023629, R01AG15928, and R01AG20098 from the NIA. FHS research is supported by NHLBI contracts N01-HC-25195 and HHSN268201500001I. This study was also supported by additional grants from the NIA (R01s AG054076, AG049607 and AG033040 and NINDS (R01 NS017950). The ERF study as a part of EUROSPAN (European Special Populations Research Network) was supported by European Commission FP6 STRP grant number 018947 (LSHG-CT-2006-01947) and also received funding from the European Community’s Seventh Framework Programme (FP7/2007-2013)/grant agreement HEALTH-F4-2007-201413 by the European Commission under the programme “Quality of Life and Management of the Living Resources” of 5th Framework Programme (no. QLG2-CT-2002-01254). High-throughput analysis of the ERF data was supported by a joint grant from the Netherlands Organization for Scientific Research and the Russian Foundation for Basic Research (NWO-RFBR 047.017.043). The Rotterdam Study is funded by Erasmus Medical Center and Erasmus University, Rotterdam, the Netherlands Organization for Health Research and Development (ZonMw), the Research Institute for Diseases in the Elderly (RIDE), the Ministry of Education, Culture and Science, the Ministry for Health, Welfare and Sports, the European Commission (DG XII), and the municipality of Rotterdam. Genetic data sets are also supported by the Netherlands Organization of Scientific Research NWO Investments (175.010.2005.011, 911-03-012), the Genetic Laboratory of the Department of Internal Medicine, Erasmus MC, the Research Institute for Diseases in the Elderly (014-93-015; RIDE2), and the Netherlands Genomics Initiative (NGI)/Netherlands Organization for Scientific Research (NWO) Netherlands Consortium for Healthy Aging (NCHA), project 050-060-810. All studies are grateful to their participants, faculty and staff. The content of these manuscripts is solely the responsibility of the authors and does not necessarily represent the official views of the National Institutes of Health or the U.S. Department of Health and Human Services.

The FUS cohorts include: the Alzheimer’s Disease Research Centers (ADRC) (P30 AG062429, P30 AG066468, P30 AG062421, P30 AG066509, P30 AG066514, P30 AG066530, P30 AG066507, P30 AG066444, P30 AG066518, P30 AG066512, P30 AG066462, P30 AG072979, P30 AG072972, P30 AG072976, P30 AG072975, P30 AG072978, P30 AG072977, P30 AG066519, P30 AG062677, P30 AG079280, P30 AG062422, P30 AG066511, P30 AG072946, P30 AG062715, P30 AG072973, P30 AG066506, P30 AG066508, P30 AG066515, P30 AG072947, P30 AG072931, P30 AG066546, P20 AG068024, P20 AG068053, P20 AG068077, P20 AG068082, P30 AG072958, P30 AG072959), Alzheimer’s Disease Neuroimaging Initiative (ADNI) (U19AG024904), Amish Protective Variant Study (RF1AG058066), Cache County Study (R01AG11380, R01AG031272, R01AG21136, RF1AG054052), Case Western Reserve University Brain Bank (CWRUBB) (P50AG008012), Case Western Reserve University Rapid Decline (CWRURD) (RF1AG058267, NU38CK000480), CubanAmerican Alzheimer’s Disease Initiative (CuAADI) (3U01AG052410), Estudio Familiar de Influencia Genetica en Alzheimer (EFIGA) (5R37AG015473, RF1AG015473, R56AG051876), Genetic and Environmental Risk Factors for Alzheimer Disease Among African Americans Study (GenerAAtions) (2R01AG09029, R01AG025259, 2R01AG048927), Gwangju Alzheimer and Related Dementias Study (GARD) (U01AG062602), Hillblom Aging Network (2014-A-004-NET, R01AG032289, R01AG048234), Hussman Institute for Human Genomics Brain Bank (HIHGBB) (R01AG027944, Alzheimer’s Association “Identification of Rare Variants in Alzheimer Disease”), Ibadan Study of Aging (IBADAN) (5R01AG009956), Longevity Genes Project (LGP) and LonGenity (R01AG042188, R01AG044829, R01AG046949, R01AG057909, R01AG061155, P30AG038072), Mexican Health and Aging Study (MHAS) (R01AG018016), Multi-Institutional Research in Alzheimer’s Genetic Epidemiology (MIRAGE) (2R01AG09029, R01AG025259, 2R01AG048927), Northern Manhattan Study (NOMAS) (R01NS29993), Peru Alzheimer’s Disease Initiative (PeADI) (RF1AG054074), Puerto Rican 1066 (PR1066) (Wellcome Trust (GR066133/GR080002), European Research Council (340755)), Puerto Rican Alzheimer Disease Initiative (PRADI) (RF1AG054074), Reasons for Geographic and Racial Differences in Stroke (REGARDS) (U01NS041588), Research in African American Alzheimer Disease Initiative (REAAADI) (U01AG052410), the Religious Orders Study (ROS) (P30 AG10161, P30 AG72975, R01 AG15819, R01 AG42210), the RUSH Memory and Aging Project (MAP) (R01 AG017917, R01 AG42210Stanford Extreme Phenotypes in AD (R01AG060747), University of Miami Brain Endowment Bank (MBB), University of Miami/Case Western/North Carolina A&T African American (UM/CASE/NCAT) (U01AG052410, R01AG028786), Wisconsin Registry for Alzheimer’s Prevention (WRAP) (R01AG027161 and R01AG054047), Mexico-Southern California Autosomal Dominant Alzheimer’s Disease Consortium (R01AG069013), Center for Cognitive Neuroscience and Aging (R01AG047649), and the A4 Study (R01AG063689, U19AG010483 and U24AG057437).

The four LSACs are: the Human Genome Sequencing Center at the Baylor College of Medicine (U54 HG003273), the Broad Institute Genome Center (U54HG003067), The American Genome Center at the Uniformed Services University of the Health Sciences (U01AG057659), and the Washington University Genome Institute (U54HG003079). Genotyping and sequencing for the ADSP FUS is also conducted at John P. Hussman Institute for Human Genomics (HIHG) Center for Genome Technology (CGT).

Biological samples and associated phenotypic data used in primary data analyses were stored at Study Investigators institutions, and at the National Centralized Repository for Alzheimer’s Disease and Related Dementias (NCRAD, U24AG021886) at Indiana University funded by NIA. Associated Phenotypic Data used in primary and secondary data analyses were provided by Study Investigators, the NIA funded Alzheimer’s Disease Centers (ADCs), and the National Alzheimer’s Coordinating Center (NACC, U24AG072122) and the National Institute on Aging Genetics of Alzheimer’s Disease Data Storage Site (NIAGADS, U24AG041689) at the University of Pennsylvania, funded by NIA. Harmonized phenotypes were provided by the ADSP Phenotype Harmonization Consortium (ADSP-PHC), funded by NIA (U24 AG074855, U01 AG068057 and R01 AG059716) and Ultrascale Machine Learning to Empower Discovery in Alzheimer’s Disease Biobanks (AI4AD, U01 AG068057). This research was supported in part by the Intramural Research Program of the National Institutes of health, National Library of Medicine. Contributors to the Genetic Analysis Data included Study Investigators on projects that were individually funded by NIA, and other NIH institutes, and by private U.S. organizations, or foreign governmental or nongovernmental organizations.

The ADSP Phenotype Harmonization Consortium (ADSP-PHC) is funded by NIA (U24 AG074855, U01 AG068057 and R01 AG059716). The harmonized cohorts within the ADSP-PHC include: the Anti-Amyloid Treatment in Asymptomatic Alzheimer’s study (A4 Study), a secondary prevention trial in preclinical Alzheimer’s disease, aiming to slow cognitive decline associated with brain amyloid accumulation in clinically normal older individuals. The A4 Study is funded by a public-private-philanthropic partnership, including funding from the National Institutes of Health-National Institute on Aging, Eli Lilly and Company, Alzheimer’s Association, Accelerating Medicines Partnership, GHR Foundation, an anonymous foundation and additional private donors, with in-kind support from Avid and Cogstate. The companion observational Longitudinal Evaluation of Amyloid Risk and Neurodegeneration (LEARN) Study is funded by the Alzheimer’s Association and GHR Foundation. The A4 and LEARN Studies are led by Dr. Reisa Sperling at Brigham and Women’s Hospital, Harvard Medical School and Dr. Paul Aisen at the Alzheimer’s Therapeutic Research Institute (ATRI), University of Southern California. The A4 and LEARN Studies are coordinated by ATRI at the University of Southern California, and the data are made available through the Laboratory for Neuro Imaging at the University of Southern California. The participants screening for the A4 Study provided permission to share their de-identified data in order to advance the quest to find a successful treatment for Alzheimer’s disease. We would like to acknowledge the dedication of all the participants, the site personnel, and all of the partnership team members who continue to make the A4 and LEARN Studies possible. The complete A4 Study Team list is available on: a4study.org/a4-study-team.; the Adult Changes in Thought study (ACT), U01 AG006781, U19 AG066567; Alzheimer’s Disease Neuroimaging Initiative (ADNI): Data collection and sharing for this project was funded by the Alzheimer’s Disease Neuroimaging Initiative (ADNI) (National Institutes of Health Grant U01 AG024904) and DOD ADNI (Department of Defense award number W81XWH-12-2-0012). ADNI is funded by the National Institute on Aging, the National Institute of Biomedical Imaging and Bioengineering, and through generous contributions from the following: AbbVie, Alzheimer’s Association; Alzheimer’s Drug Discovery Foundation; Araclon Biotech; BioClinica, Inc.; Biogen; Bristol-Myers Squibb Company; CereSpir, Inc.; Cogstate; Eisai Inc.; Elan Pharmaceuticals, Inc.; Eli Lilly and Company; EuroImmun; F. Hoffmann-La Roche Ltd and its affiliated company Genentech, Inc.; Fujirebio; GE Healthcare; IXICO Ltd.;Janssen Alzheimer Immunotherapy Research & Development, LLC.; Johnson & Johnson Pharmaceutical Research & Development LLC.; Lumosity; Lundbeck; Merck & Co., Inc.;Meso Scale Diagnostics, LLC.; NeuroRx Research; Neurotrack Technologies; Novartis Pharmaceuticals Corporation; Pfizer Inc.; Piramal Imaging; Servier; Takeda Pharmaceutical Company; and Transition Therapeutics. The Canadian Institutes of Health Research is providing funds to support ADNI clinical sites in Canada. Private sector contributions are facilitated by the Foundation for the National Institutes of Health (www.fnih.org). The grantee organization is the Northern California Institute for Research and Education, and the study is coordinated by the Alzheimer’s Therapeutic Research Institute at the University of Southern California. ADNI data are disseminated by the Laboratory for Neuro Imaging at the University of Southern California; Estudio Familiar de Influencia Genetica en Alzheimer (EFIGA): 5R37AG015473, RF1AG015473, R56AG051876; the Health & Aging Brain Study – Health Disparities (HABS-HD), supported by the National Institute on Aging of the National Institutes of Health under Award Numbers R01AG054073, R01AG058533, R01AG070862, P41EB015922, and U19AG078109; the Korean Brain Aging Study for the Early Diagnosis and Prediction of Alzheimer’s disease (KBASE), which was supported by a grant from Ministry of Science, ICT and Future Planning (Grant No: NRF-2014M3C7A1046042); Memory & Aging Project at Knight Alzheimer’s Disease Research Center (MAP at Knight ADRC): The Memory and Aging Project at the Knight-ADRC (Knight-ADRC). This work was supported by the National Institutes of Health (NIH) grants R01AG064614, R01AG044546, RF1AG053303, RF1AG058501, U01AG058922 and R01AG064877 to Carlos Cruchaga. The recruitment and clinical characterization of research participants at Washington University was supported by NIH grants P30AG066444, P01AG03991, and P01AG026276. Data collection and sharing for this project was supported by NIH grants RF1AG054080, P30AG066462, R01AG064614 and U01AG052410. We thank the contributors who collected samples used in this study, as well as patients and their families, whose help and participation made this work possible. This work was supported by access to equipment made possible by the Hope Center for Neurological Disorders, the Neurogenomics and Informatics Center (NGI: https://neurogenomics.wustl.edu/) and the Departments of Neurology and Psychiatry at Washington University School of Medicine; National Alzheimer’s Coordinating Center (NACC): The NACC database is funded by NIA/NIH Grant U24 AG072122. SCAN is a multi-institutional project that was funded as a U24 grant (AG067418) by the National Institute on Aging in May 2020. Data collected by SCAN and shared by NACC are contributed by the NIA-funded ADRCs as follows: P30 AG062429 (PI James Brewer, MD, PhD), P30 AG066468 (PI Oscar Lopez, MD), P30 AG062421 (PI Bradley Hyman, MD, PhD), P30 AG066509 (PI Thomas Grabowski, MD), P30 AG066514 (PI Mary Sano, PhD), P30 AG066530 (PI Helena Chui, MD), P30 AG066507 (PI Marilyn Albert, PhD), P30 AG066444 (PI John Morris, MD), P30 AG066518 (PI Jeffrey Kaye, MD), P30 AG066512 (PI Thomas Wisniewski, MD), P30 AG066462 (PI Scott Small, MD), P30 AG072979 (PI David Wolk, MD), P30 AG072972 (PI Charles DeCarli, MD), P30 AG072976 (PI Andrew Saykin, PsyD), P30 AG072975 (PI David Bennett, MD), P30 AG072978 (PI Neil Kowall, MD), P30 AG072977 (PI Robert Vassar, PhD), P30 AG066519 (PI Frank LaFerla, PhD), P30 AG062677 (PI Ronald Petersen, MD, PhD), P30 AG079280 (PI Eric Reiman, MD), P30 AG062422 (PI Gil Rabinovici, MD), P30 AG066511 (PI Allan Levey, MD, PhD), P30 AG072946 (PI Linda Van Eldik, PhD), P30 AG062715 (PI Sanjay Asthana, MD, FRCP), P30 AG072973 (PI Russell Swerdlow, MD), P30 AG066506 (PI Todd Golde, MD, PhD), P30 AG066508 (PI Stephen Strittmatter, MD, PhD), P30 AG066515 (PI Victor Henderson, MD, MS), P30 AG072947 (PI Suzanne Craft, PhD), P30 AG072931 (PI Henry Paulson, MD, PhD), P30 AG066546 (PI Sudha Seshadri, MD), P20 AG068024 (PI Erik Roberson, MD, PhD), P20 AG068053 (PI Justin Miller, PhD), P20 AG068077 (PI Gary Rosenberg, MD), P20 AG068082 (PI Angela Jefferson, PhD), P30 AG072958 (PI Heather Whitson, MD), P30 AG072959 (PI James Leverenz, MD); National Institute on Aging Alzheimer’s Disease Family Based Study (NIA-AD FBS): U24 AG056270; Religious Orders Study (ROS): P30AG10161,R01AG15819, R01AG42210; Memory and Aging Project (MAP - Rush): R01AG017917, R01AG42210; Minority Aging Research Study (MARS): R01AG22018, R01AG42210; the Texas Alzheimer’s Research and Care Consortium (TARCC), funded by the Darrell K Royal Texas Alzheimer’s Initiative, directed by the Texas Council on Alzheimer’s Disease and Related Disorders; Washington Heights/Inwood Columbia Aging Project (WHICAP): RF1 AG054023;and Wisconsin Registry for Alzheimer’s Prevention (WRAP): R01AG027161 and R01AG054047. Additional acknowledgments include the National Institute on Aging Genetics of Alzheimer’s Disease Data Storage Site (NIAGADS, U24AG041689) at the University of Pennsylvania, funded by NIA.

